# Barriers to healthcare for autistic adults: Consequences & policy implications. A cross-sectional study

**DOI:** 10.1101/2020.04.01.20050336

**Authors:** Mary Doherty, Stuart D Neilson, Jane D O'Sullivan, Laura Carravallah, Mona Johnson, Walter Cullen, Louise Gallagher

## Abstract

**Background:** Autistic people experience significantly poorer physical and mental health along with reduced life expectancy.

**Aim:** To identify self-reported barriers to primary care by autistic adults compared to parents of autistic children and non-autistic adults and link these barriers to self-reported adverse health consequences.

**Design and Setting:** Following consultation with the autistic community at an autistic conference, Autscape, a quantitative and qualitative survey was developed.

**Method:** The self-report survey was administered online through social media platforms.

**Results:** The 57-item online survey was completed by 507 autistic adults, 196 parents of autistic children and 157 control subjects. 79.7% of autistic adults, 52.8% of parents and 36.5% of controls reported difficulty visiting a GP. The highest-rated barriers by autistic adults were deciding if symptoms warrant a GP visit (72.2%), difficulty making appointments by telephone (61.9%), not feeling understood (55.8%), difficulty communicating with their doctor (53.1%) and the waiting room environment (50.5%).

Autistic adults reported a preference for online or text based appointment booking, facility to email in advance the reason for consultation, first or last clinic appointment and a quiet place to wait.

Increased adverse health outcomes reported by autistic adults correlated with difficulty attending, including untreated physical and mental health conditions, not attending specialist referral or screening programmes, requiring more extensive treatment or surgery due to late presentations, and untreated potentially life threatening conditions.

**Conclusion:** Reduction of healthcare inequalities for autistic people requires that healthcare providers understand autistic perspectives and communication needs. Adjustments for autism specific needs are as necessary as ramps for wheelchair users.

**How this fits in:**

- Adverse health outcomes are common among autistic people and so it is important to understand how we can promote access to primary care.
- This cross sectional study indicates that 79.7% of autistic patients (compared to 36.5% of controls) reported difficulty visiting a GP.
- Common barriers were: deciding if symptoms warrant a GP visit, difficulty using the telephone to book appointments, not feeling understood and difficulty communicating with their doctor.
- Common suggestions to promote access included: online or text based appointment booking facility, emailing in advance the reason for consultation, providing first or last clinic appointment and having a quiet place to wait.

## Introduction

Autism is a common neurodevelopmental condition affecting 1-2% of the population[1]. Most autistic people are adult, do not have intellectual disability and are likely to be undiagnosed[2]. Doctors may underestimate of the number of autistic patients under their care[3,4].

Autistic adults have poor physical and mental health compared to the general population[5]. Most medical conditions are more common[6,7], including diabetes, hypertension and obesity[8]. Autistic people experience premature mortality[9,10,11]. Life expectancy is reduced by 16-30years, with increased mortality reported across almost all diagnostic categories[9] and in-hospital mortality is increased[12]. Autistic people are three times more likely to use emergency departments, to require inpatient admission, and to die after attending emergency care[13].

Alongside increased health needs, autistic people report a greater likelihood that their needs are unmet[14]. Pervasive, multifactorial barriers to healthcare access are experienced[15]. Some are shared by other disabled people, but autistic patients experience additional autism-specific barriers[16]. A recent systematic review identified patient-provider communication, sensory sensitivities, and executive functioning/planning difficulties as important barriers to healthcare[17]. Negative experiences with healthcare providers was separately identified as a prominent barrier[18].

In response to primary legislation[19] and statutory guidance[20] The Royal College of General Practitioners (RCGP) developed an Autism Patient Charter[21] that recommended: staff awareness and training; autism friendly environment; reasonable adjustments following disclosure or clinical suspicion of autism; patient-tailored communications; and behaviour-sensitive accommodations. In spite of efforts to champion autism, proposals to formalise autism training[18,22] and specific awareness-raising interventions[21], almost 40% of GPs report no formal training in autism and have limited confidence in managing autistic patients[22]. Greater autism awareness exists where GPs have personal knowledge of autism, either through a relative or friend on the autistic spectrum, or because they themselves are autistic[22].

Communication skills training for health care providers is the most pressing need[4]. GPs[22] and hospital specialists[3] self-report difficulties communicating with autistic patients. Only 25% of primary healthcare providers reported high confidence in communicating with autistic adult patients, or identifying and making necessary accommodations[4].

This study aimed to identify self-reported barriers to primary health care faced by autistic adults with a focus on autism-specific communication, sensory and procedural considerations. Self-reported consequences were captured to add a narrative frame to the existing evidence base around disparities in health outcomes. Since non-autistic parents of autistic children experience pronounced increase in morbidity and mortality related to cancer and vascular diseases compared to parents of non-autistic children, this paper further illuminates the barriers experienced by that group[23]. This is to our knowledge the largest study of primary healthcare barriers to date and benefits from a high degree of participatory design by the autistic community making a unique and substantial contribution to the current literature.

## Methods

An initial survey was undertaken in August 2018 at Autscape, an annual 3-day residential conference organized by and for autistic people[24]. This was undertaken as a quality improvement project for an “Autism Friendly Town” initiative by AsIAm, Ireland’s National Autism Charity[25,26] to inform autism training for local healthcare providers. A 16-item questionnaire, entitled “What do you wish your GP knew about autism?” was distributed in paper and online formats containing open-ended questions and open-text response fields, allowing for qualitative answers. Participants at Autscape included autistic attendees, non-autistic family members and carers, who were capable of completing the survey. The majority of Autscape attendees typically have low to moderate support needs. Seventy-five responses were received, reviewed (by MD) and grouped thematically under broad themes. Thematic analysis was pragmatic, based on face validity rather than a formalised qualitative analysis.

Next a 57-item online survey was developed to investigate barriers to primary care in a larger sample of autistic adults and compare with parents of autistic children and a non-autistic adult control group.

The survey was adapted for 3 groups of respondents: 1. Autistic adults, including those who were parents (Autistic); 2. Parents of autistic children, who did not identify as autistic themselves (Parent); 3. Non autistic adults who did not have an autistic child (Control). All were asked to respond specifically about seeking healthcare for themselves, not for their children. Respondents were directed to the appropriate survey however a small number(n=55) answered the incorrect survey. As the questions were the same it was possible to assign their survey responses to the appropriate group.

Question types included yes-no responses, single- and multiple-item selections from a list, Likert scales, and open-ended short answer. Questions explored the specific barriers encountered and the consequences of such barriers, in particular the reasons for delaying or avoiding a visit, difficulties booking, planning and waiting for a GP visit, challenges during a consultation, including communication, sensory, emotional and organisation issues as well as social supports available. Reasonable adjustments to standard care and access were also explored as well as self-reported consequences of failure to access healthcare.

Participants were recruited through social media, primarily Twitter and Facebook, via both personal pages and in autism related groups and shared on the AsIAm website.

Survey data were first collected between October 2018 and April 2019. Reminders were given by repeat posting on social media platforms. Preliminary analysis revealed a recurring theme of total non-engagement with healthcare providers, despite expressed healthcare needs. Consequently the survey was altered to add response options “I do not attend any medical practice?” and “I do not attend any doctor” to the questions “Do you usually attend the same medical practice?” and “Do you usually see the same doctor?”. The amended survey was run again over a two-week period in August 2019. Due to unquantifiable duplication of responses between the first and second data collection periods, only the data from the second collection period are presented here. Analysis revealed good correlation between both sets of results.

Significance of between-group differences on categorical variables was assessed using 2×2 or 2xk Chi-squared test of differences between paired groups: autistic adults with control participants, autistic adults with parents, and parents with control participants. A Wilcoxon-Mann-Whitney U test was similarly employed on ordinal scales.

## Results

### Participants

860 respondents completed the online survey: 507 autistic adults, 196 non-autistic parents of autistic children and 157 ‘control’ adults (Table 1).

**Table 1:**
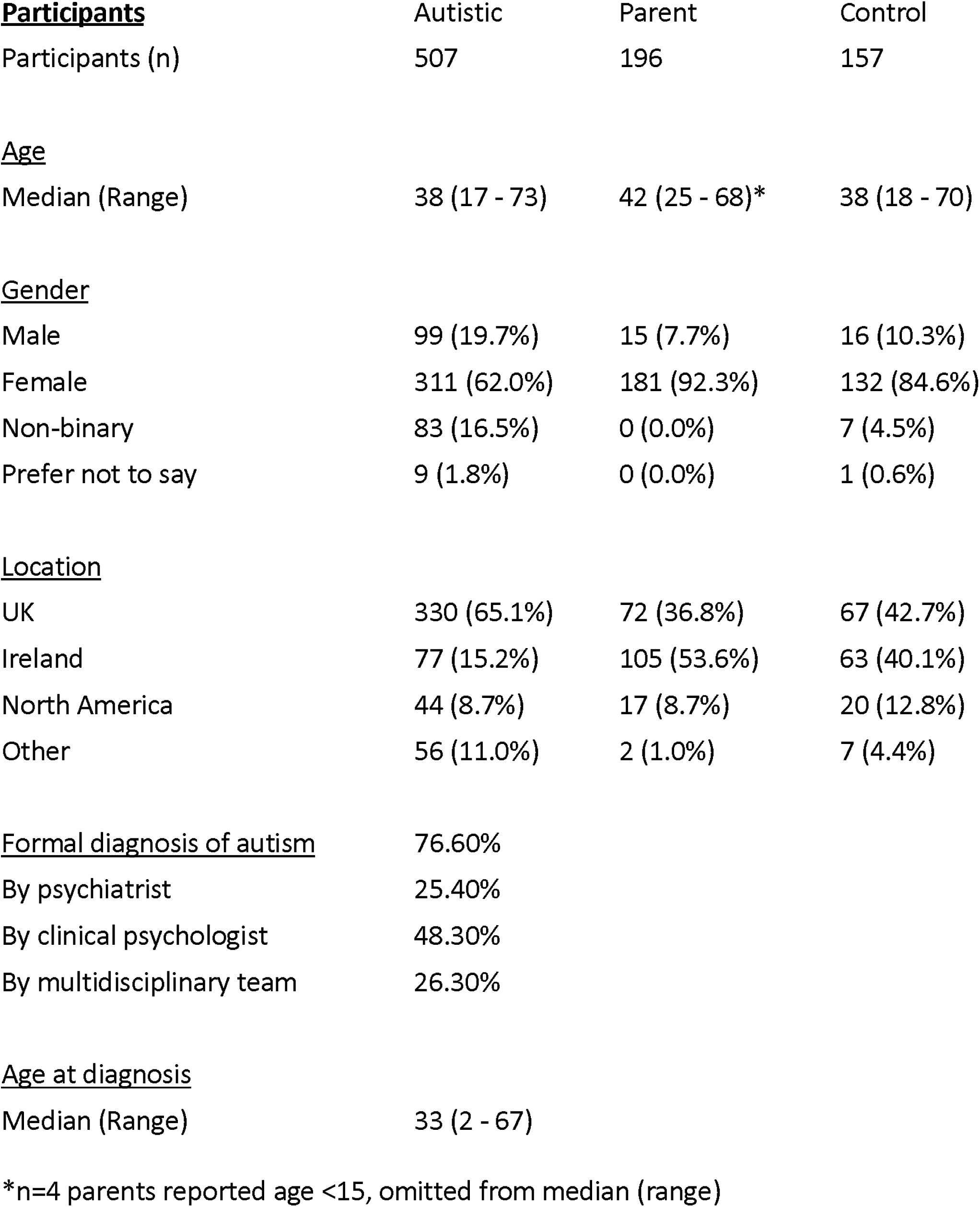
Participant Characteristics

### Barriers to access

79.7% of autistic respondents reported difficulty visiting a GP when needed, compared to 52.8% of parents and 36.5% of controls. See Table 2 for specific barriers.

**Table 2:** Access Barriers to Healthcare

| **<u>Access Barriers</u>** | **Autistic** | **Parent** | **Control** | **Autistic.Control** | **Autistic.Parent** | **Parent.Control** |
| --- | --- | --- | --- | --- | --- | --- |
| Difficulty attending a GP | 400 (79.7%) | 103 (52.8%) | 57 (36.5%) | p<0.001** | p<0.001** | p<0.001** |
| **<u>Which of the following would cause you to delay or avoid seeing your doctor when you need to?</u>** |  |  |  |  |  |  |
|  | **Autistic** | **Parent** | **Control** | **Autistic.Control** | **Autistic.Parent** | **Parent.Control** |
| Difficulty deciding if symptoms warrant a GP visit | 366 (72.2%) | 82 (41.8%) | 102 (65.0%) | p=0.102 ns | p<0.001** | p<0.001** |
| Difficulty using the telephone to book appointment | 314 (61.9%) | 10 (5.1%) | 25 (15.9%) | p<0.001** | p<0.001** | p<0.001** |
| Not feeling understood | 283 (55.8%) | 29 (14.8%) | 21 (13.4%) | p<0.001** | p<0.001** | p=0.821 ns |
| Difficulty communicating with the doctor during the appointment | 269 (53.1%) | 13 (6.6%) | 10 (6.4%) | p<0.001** | p<0.001** | p=1.000 ns |
| The waiting room environment | 256 (50.5%) | 25 (12.8%) | 12 (7.6%) | p<0.001** | p<0.001** | p=0.167 ns |
| Long wait to get an appointment | 251 (49.5%) | 81 (41.3%) | 70 (44.6%) | p=0.324 ns | p=0.062 ns | p=0.612 ns |
| Difficulty planning an appointment in advance | 243 (47.9%) | 59 (30.1%) | 51 (32.5%) | p<0.001** | p<0.001** | p=0.715 ns |
| Inability to see a known or preferred doctor | 241 (47.5%) | 42 (21.4%) | 35 (22.3%) | p<0.001** | p<0.001** | p=0.948 ns |
| Difficulty communicating with the reception staff | 235 (46.4%) | 11 (5.6%) | 13 (8.3%) | p<0.001** | p<0.001** | p=0.437 ns |
| Not having enough time to visit the doctor | 174 (34.3%) | 69 (35.2%) | 61 (38.9%) | p=0.346 ns | p=0.894 ns | p=0.552 ns |
| No online booking system | 160 (31.6%) | 32 (16.3%) | 35 (22.3%) | p=0.033* | p<0.001** | p=0.199 ns |
| Waiting to see the doctor is too difficult | 114 (22.5%) | 16 (8.2%) | 7 (4.5%) | p<0.001** | p<0.001** | p=0.236 ns |
| Needing a support person to come with me | 106 (20.9%) | 11 (5.6%) | 7 (4.5%) | p<0.001** | p<0.001** | p=0.806 ns |
| There is an online booking system but it's confusing | 102 (20.1%) | 11 (5.6%) | 9 (5.7%) | p<0.001** | p<0.001** | p=1.000 ns |
| Not having anyone to look after my child | 66 (13.0%) | 73 (37.2%) | 18 (11.5%) | p=0.708 ns | p<0.001** | p<0.001** |
| None of the above | 6 (1.2%) | 16 (8.2%) | 17 (10.8%) | p<0.001** | p<0.001** | p=0.502 ns |
| **<u>Regular GP</u>** |  |  |  |  |  |  |
|  | **Autistic** | **Parent** | **Control** | **Autistic.Control** | **Autistic.Parent** | **Parent.Control** |
| Usually attend the same medical practice | 467 (92.7%) | 185 (94.4%) | 144 (92.3%) | p=0.908 | p=0.594 | p=0.718 |
| Usually see the same doctor | 280 (55.3%) | 123 (62.8%) | 74 (47.4%) | p=0.223 | p=0.138 | p=0.012* |
| I don't attend any doctor | 20 (4.0%) | 4 (2.0%) | 7 (4.5%) | p=0.223 | p=0.138 | p=0.012* |

Table 2: Access Barriers to Healthcare (contd)
| <u>Usual reason for visit</u> | **Autistic** | **Parent** | **Control** | **Autistic.Control** | **Autistic.Parent** | **Parent.Control** |
| --- | --- | --- | --- | --- | --- | --- |
| Physical condition or illness | 435 (85.8%) | 173 (88.3%) | 145 (92.4%) | p=0.043 * | p=0.462 ns | p=0.272 ns |
| Mental health difficulties | 310 (61.1%) | 85 (43.4%) | 43 (27.4%) | p<0.001 ** | p<0.001 ** | p=0.003 ** |
| Issues directly related to autism | 113 (22.3%) | 41 (20.9%) | 0 (0.0%) | p<0.001 ** | p=0.770 ns | p<0.001 ** |

### Communication

Factors causing an autistic patient to delay or avoid seeing their doctor when required included: difficulty using the telephone to book an appointment (61.9%); not feeling understood (55.8%); difficulty communicating with the doctor (53.1%) and with reception staff (46.4%). 59% of autistic respondents reported difficulty communicating during a consultation “all the time” or “frequently” compared to 11.3% of parents and 11.5% of controls (p<0.001).

Autistic respondents reported avoiding the telephone (77.9%), voicemail (61.3%) and face-to-face verbal communication (30.0%). 41.0% reported that it is “easier for me to communicate in writing” (Table 3).

**Table 3:** Communication Barriers

| **<u>Reasons to avoid or delay GP visit (Communication)</u>** | **Autistic** | **Parent** | **Control** | **Autistic.Control** | **Autistic.Parent** | **Parent.Control** |
| --- | --- | --- | --- | --- | --- | --- |
| Difficulty using the telephone to book an appointment | 314 (61.9%) | 10 (5.1%) | 25 (15.9%) | p<0.001** | p<0.001** | p=0.001** |
| Not feeling understood | 283 (55.8%) | 29 (14.8%) | 21 (13.4%) | p<0.001** | p<0.001** | p=0.821 ns |
| Difficulty communicating with the doctor during the appointment | 269 (53.1%) | 13 (6.6%) | 10 (6.4%) | p<0.001** | p<0.001** | p=1.000 ns |
| Difficulty communicating with the reception staff | 235 (46.4%) | 11 (5.6%) | 13 (8.3%) | p<0.001** | p<0.001** | p=0.437 ns |
| **<u>Communication preferences</u>** |  |  |  |  |  |  |
| Telephone generally avoided where possible | 395 (77.9%) | 60 (30.6%) | 59 (37.6%) | p<0.001** | p<0.001** | p=0.207 ns |
| Voicemail generally avoided where possible | 311 (61.3%) | 54 (27.6%) | 61 (38.9%) | p<0.001** | p<0.001** | p=0.033* |
| Verbal, face-to-face communication generally avoided where possible | 152 (30.0%) | 31 (15.8%) | 12 (7.6%) | p<0.001** | p<0.001** | p=0.03* |
| It is easier for me to communicate in writing | 208 (41.0%) | 11 (5.6%) | 13 (8.3%) | p<0.001** | p<0.001** | p=0.437 ns |
| **<u>Communication challenges</u>** |  |  |  |  |  |  |
| Anxiety makes it harder to communicate | 395 (77.9%) | 49 (25.0%) | 42 (26.8%) | p<0.001** | p<0.001** | p=0.801 ns |
| I have difficulty prioritising when describing medical symptoms | 333 (65.7%) | 38 (19.4%) | 34 (21.7%) | p<0.001** | p<0.001** | p=0.695 ns |
| I need to give the whole story and not leave anything out | 332 (65.5%) | 30 (15.3%) | 18 (11.5%) | p<0.001** | p<0.001** | p=0.373 ns |
| I need extra time to process what is being said | 286 (56.4%) | 18 (9.2%) | 12 (7.6%) | p<0.001** | p<0.001** | p=0.746 ns |
| I can't describe my pain or symptoms accurately | 272 (53.6%) | 26 (13.3%) | 36 (22.9%) | p<0.001** | p<0.001** | p=0.026* |
| Verbal communication is difficult | 234 (46.2%) | 8 (4.1%) | 11 (7.0%) | p<0.001** | p<0.001** | p=0.331 ns |
| I express emotions differently e.g. I can appear angry when I am afraid or in pain | 227 (44.8%) | 14 (7.1%) | 6 (3.8%) | p<0.001** | p<0.001** | p=0.267 ns |
| Sensory issues make communication more difficult | 156 (30.8%) | 10 (5.1%) | 2 (1.3%) | p<0.001** | p<0.001** | p=0.094 ns |
| None of the above (Communication) | 11 (2.2%) | 59 (30.1%) | 58 (36.9%) | p<0.001** | p<0.001** | p=0.214 ns |

### Predictability & control

Autistic respondents reported difficulty not knowing the wait duration (70.0%), what would happen during the consultation (63.1%), which doctor they would see (57.6%) and the consultation length (39.6%). Parents had difficulty with unknown waiting time (49.0%) (Supplementary Table 1).

### Sensory processing

The waiting room environment was a barrier for 50.5% of autistic respondents. Sensory issues make communication more difficult for 30.8% of the autistic group (Table 2). Only 10.1% of autistic respondents marked “none of the above” to sensory questions compared to 62.2% of parents and 71.3% of controls (Table 4).

**Table 4:** Sensory Barriers

| **<u>Reasons to avoid or delay GP visit (Sensory)</u>** | **Autistic** | **Parent** | **Control** | **Autistic.Control** | **Autistic.Parent** | **Parent.Control** |
| --- | --- | --- | --- | --- | --- | --- |
| The waiting room environment | 256 (50.5%) | 25 (12.8%) | 12 (7.6%) | p<0.001 ** | p<0.001 ** | p=0.167 ns |
| **<u>Specific sensory challenges</u>** |  |  |  |  |  |  |
| Noise in the waiting room from other patients | 319 (62.9%) | 37 (18.9%) | 19 (12.1%) | p<0.001 ** | p<0.001 ** | p=0.113 ns |
| Bright or fluorescent lights | 268 (52.9%) | 22 (11.2%) | 14 (8.9%) | p<0.001 ** | p<0.001 ** | p=0.593 ns |
| Crowded waiting area | 299 (59.0%) | 47 (24.0%) | 22 (14.0%) | p<0.001 ** | p<0.001 ** | p=0.027 * |
| Uncomfortable furniture | 195 (38.5%) | 21 (10.7%) | 11 (7.0%) | p<0.001 ** | p<0.001 ** | p=0.308 ns |
| Unexpected touch | 193 (38.1%) | 12 (6.1%) | 9 (5.7%) | p<0.001 ** | p<0.001 ** | p=1.000 ns |
| Music playing in the waiting room | 172 (33.9%) | 12 (6.1%) | 9 (5.7%) | p<0.001 ** | p<0.001 ** | p=1.000 ns |
| Smells in the waiting room | 171 (33.7%) | 26 (13.3%) | 8 (5.1%) | p<0.001 ** | p<0.001 ** | p=0.016 * |
| Touch, such as during examination | 160 (31.6%) | 15 (7.7%) | 11 (7.0%) | p<0.001 ** | p<0.001 ** | p=0.979 ns |
| Noise from the reception desk | 140 (27.6%) | 11 (5.6%) | 4 (2.5%) | p<0.001 ** | p<0.001 ** | p=0.249 ns |
| Smells in the doctor's office | 104 (20.5%) | 10 (5.1%) | 6 (3.8%) | p<0.001 ** | p<0.001 ** | p=0.751 ns |
| None of the above (Sensory) | 51 (10.1%) | 122 (62.2%) | 112 (71.3%) | p<0.001 ** | p<0.001 ** | p=0.092 ns |

### Judgement & attitudes

Autistic people experience more anxiety accessing healthcare; only 3.4% stated they didn’t feel anxious going to the doctor, compared to 26.6% of parents and 33.1% of controls. Autistic patients report being “concerned I won’t be taken seriously when I describe my symptoms” (67.3%); worried about “wasting the doctor’s time” (65.7%) and “being considered a hypochondriac” (65.1%): experience difficulty “asking for help” (62.7%) and “discussing mental health” (59.4%) (Supplementary Table 2).

### Disclosure of diagnosis

Only 61.5% of autistic patients reported their doctor knew they were autistic, 22.4% were unsure and 16.1% hadn’t disclosed their diagnosis. In contrast, 88.3% of parents reported their doctor knew their child’s diagnosis.

### Planning & Organising

Autistic respondents reported greater difficulties with organisation and planning for healthcare, including difficulties “making an appointment in advance” (59.2%), “prioritising my health issues” (58.4%) and “making changes to my lifestyle or habits” (55.6%). 45.4% reported forgetting a medical appointment and 29.8% attended on the wrong day. Parents (41.8%) reported significantly less difficulty deciding if symptoms warrant a GP visit than autistic (72.2%) and control (65.0%) respondents. (Supplementary Table 3).

### Support needs

Autistic adults more frequently reported physical mobility needs (15.9%), and unmet support needs such as “needing a support person to come with me” (20.9%). This extended to secondary care: 17.4% had no one to support hospital visits, collection from hospital (19.5%), or home care following discharge (25.8%). “Not having anyone to look after my child” was a greater barrier for parents (37.2%) than autistic (13.0%) or control (11.5%) groups (Supplementary Table 4).

### Adverse consequences

Autistic respondents reported adverse consequences of these barriers more frequently than parents or controls, including untreated mental (69.0%) and physical (62.8%) health conditions or “did not attend referral to a specialist” (47.1%). Notably a large proportion were told they “should have seen a doctor sooner” (59.9%), “required more extensive treatment or surgery” (35.9%), or did not access treatment for a “potentially serious or life threatening condition” (34.2%). Additionally, they were significantly less likely to “attend on schedule for screening programmes” than the other two groups (Figure 1).

**Figure 1.**
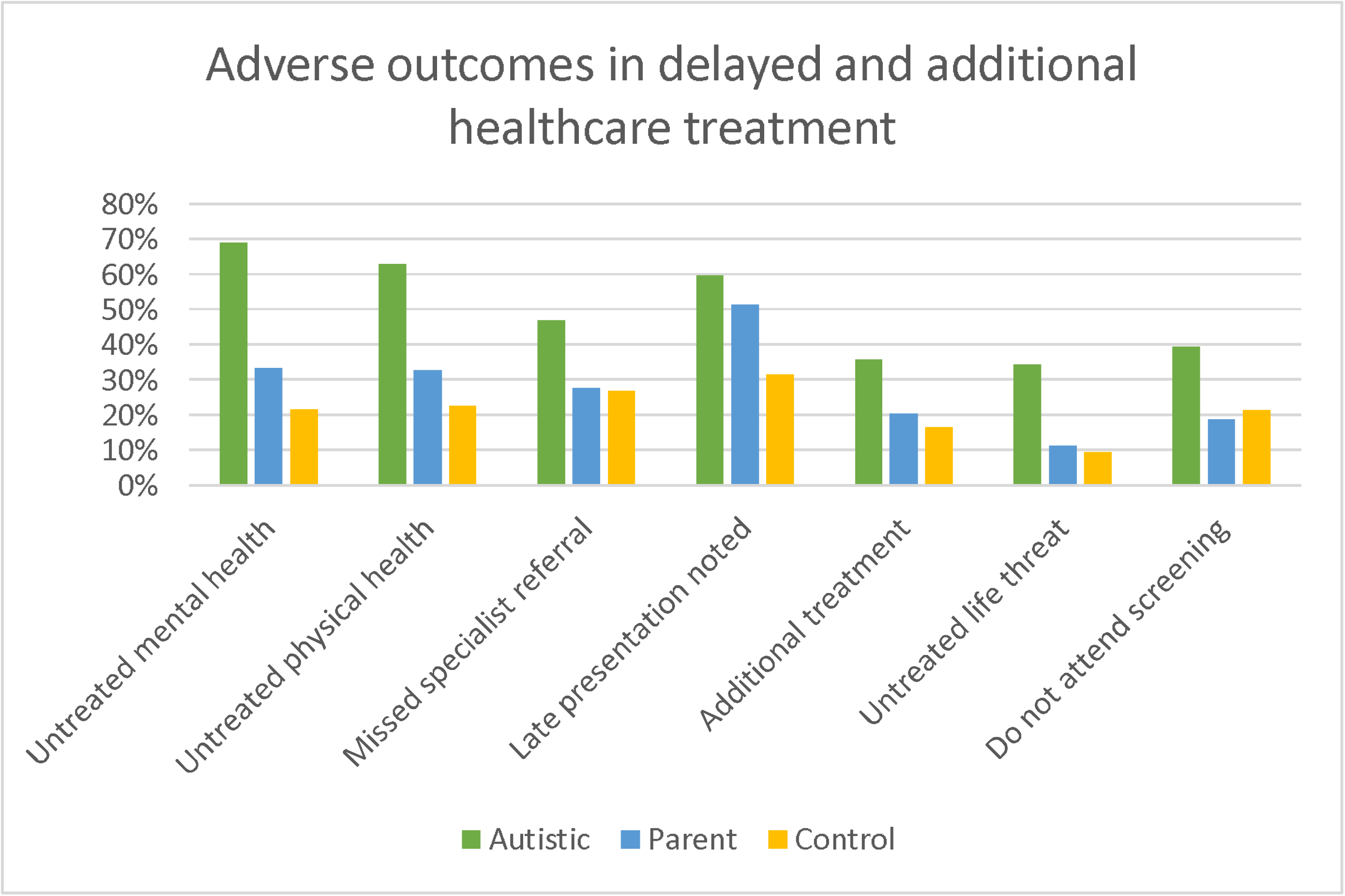
Adverse Healthcare Outcomes Late presentation noted for both autistic & parent group, compared to control (p<0.001) For all other comparisons between autistic group and parents or controls p<0.001.

Compared to autistic respondents who had no difficulty visiting a doctor, those who experienced difficulty reported more untreated mental and physical health conditions (p<0.001), not attending specialist referral (p<0.001), needing more extensive treatment (p=0.009), untreated life-threatening conditions (p=0.006) and not attending screening (p=0.028). The autistic respondents (4.0%) and parents (2.0%) who do not attend any doctor differed from the controls (4.5%) in two areas: all had difficulty visiting the doctor when needed, compared to 50% of controls (p=0.002). 95% of autistic non-attenders and 75% of parent non-attenders have experienced at least one delayed treatment outcome, compared to 43% of non-attending controls (p=0.01). There were no significant differences in difficulty attending, barriers experienced or adverse outcomes between formally diagnosed autistic respondents and those who self-identified as autistic.

### Facilitators

While most respondents reported booking an appointment online would facilitate access, autistic patients specified a need to “email my doctor in advance with a description of the issue I need to discuss” (62.3%), “wait in a quiet place or outside until my turn” (56.0%), and “book an appointment by text” (41.2%). Autistic patients and parents would be helped if they “could book the first or last appointment” (41.4% and 40.3%) and had a “sensory box available in the waiting room” (15.8% and 14.3%) (Supplementary Table 5).

Despite the outlined difficulties of visiting their doctor, autistic patients and parents felt their relationship with their GP was “very important” or “important” significantly more than controls (69.9% autistic and 76.5% parent vs. 56.1% control, p=0.001), but only 33.3% of autistic patients reported a good relationship with their doctor(p<0.001).

## Discussion

### Summary

This study describes the results of a large survey that was undertaken with a convenience sample of autistic people and compares their experiences with parents of autistic children and non-autistic adults. The study adds to the literature highlighting the barriers faced by autistic people in accessing and engaging with primary healthcare, which in our study included greater difficulties deciding when to seek care, reluctance to bother their GP, difficulties planning appointments and greater communication difficulties with particular emphasis on use of the telephone. Communication was also impaired by anxiety and sensory issues.

We linked those barriers to self-reported adverse outcomes. Our data indicated that autistic help-seeking occurs later in the natural course of an illness, with self-reported reduced attendance for screening, late presentations, missed opportunities for early detection and more extensive therapy being required. Autistic people delayed or avoided healthcare because they didn’t feel understood by their doctors. Furthermore, a significant minority of autistic adults did not disclose their autism diagnosis which further prevents the identification of their autism-specific needs. These barriers have real consequences, as evidenced in reduced life expectancy, and higher levels of physical and mental health conditions amongst autistic people.

Adverse health outcomes have also been highlighted in parents of autistic children[23]. Parents in this survey reported similar experiences to autistic adults, but reasons included prioritising their child’s needs and difficulties with childcare. The self-reported barriers and consequences lay between those reported by autistic and control respondents. Parents were more likely to disclose their child’s autism diagnosis to their GP in contrast to autistic adults. Autistic adults potentially could benefit from enhanced awareness of the need to disclose this information and the potential benefits to their care.

### Limitations

As the study used a convenience sample and self-report survey the generalisability of the data may be limited. Formal qualitative research methods were not used. Respondents required the ability to respond to the survey which means that the perspectives of autistic people with reduced ability to self-report were not represented.

### Strengths

The study arose from a community-identified need to develop autism awareness training for healthcare providers and benefits from being driven by an autistic-led research team using participatory methods, therefore it reflected the priorities of the autistic community and increased the likelihood of genuine responses being received. It provided a unique picture of autistic adults’ healthcare experiences, in particular highlighting the difficulty with using the telephone which is a distilled, concentrated essence of verbal communication. It also captured those entirely excluded from healthcare due to access barriers. This is the first comparison of the healthcare experiences of autistic adults, parents of autistic children (an under-represented group in the healthcare literature) and adult controls.

### Comparison with existing literature

This study confirms the findings of Nicolaidis[14], Raymaker[16] and several recent reviews on the topic, Mason[17], Bradshaw[27] and Walsh[15] which all showed barriers for autistic adults can be divided into three groups: (1)patient-level factors; (2)provider-level factors; and (3)system-level factors.

Our study stratifies individual barriers from the perspective of the autistic person, which addresses healthcare provider training needs identified in Nicolaidis[4]. It couples these barriers to the self-reported adverse consequences, highlighting some of the factors which may lead to excess morbidity and mortality in the autistic population.

### Implications for Research

This study suggests a need for personalised healthcare access plans. A prior study investigated using a pre-visit telephone call to identify individualised accommodations[28]. Our data suggest that this could be problematic for autistic adults. The AASPIRE Healthcare Toolkit[29] includes a publicly available online program which generates a computerised report of required healthcare accommodations. Adaptation of such a toolkit for use in NHS General Practice should be considered. Social care interventions and healthcare facilitators in general practice have shown benefit with a vulnerable population[30], similar approaches could benefit an autistic population. The significant difficulties amongst the small number of autistic people not registered with any GP indicate an urgent need for further research into this group.

### Implications for Clinical Practice

Figure 2 shows elements of an autism friendly practice that we propose. Such adjustments would minimise anxiety, manage sensory issues, and ensure mutual understanding with clear, unambiguous communication. Autism friendly practices should employ a personalised approach, with a healthcare access needs assessment and, where possible, a specialist liaison nurse or facilitator.

### Implications for Policy

Given the known barriers to care and associated impacts, the extension of annual health checks to autistic adults[31, 32] and the recently announced Oliver McGowan Mandatory Training in Learning Disability and Autism[33] are welcome. These will bring important benefits provided they are directly informed by the autistic community and autistic healthcare providers. Autism registries in GP practice have also been recommended[34,35] which would increase recognition of autism. The success of such initiatives will depend on greater awareness by medical practitioners of autistic culture and communication needs. Specific training for GPs in core training and continuing professional development would be beneficial. GPs with a special interest in autism should be facilitated to develop their skills, but management of general health needs and co-occurring conditions should be within the remit of every GP. Implementing existing autism legislation or development where lacking is urgently required to reduce health inequalities for autistic people.

## Conclusion

Significant progress towards eliminating healthcare inequalities for autistic people can only be achieved by understanding the healthcare experiences and access barriers for this vulnerable patient group in general practice.

These barriers represent not so much a failure to deliver or to avail of healthcare, but a lack of intersection between the communication patterns of autistic healthcare users and non-autistic providers, a phenomenon described as The Double Empathy Problem[36].

Reasonable accommodations are legally[37] and morally required, and adjustments for communication needs are as necessary for autistic people as ramps for wheelchair users.

### Ethical approval

Ethical approval was obtained from SJH/TUH Research Ethics Committee, Tallaght University Hospital, Dublin

## Data Availability

Data is not available.

## Acknowledgements

We acknowledge the input received from the autistic adult community recruited via local groups and online contacts during the development of the online survey. Assistance with content, structure and proofreading of the surveys was received from nine autistic adults in Ireland and the UK. We received assistance from members of peer support group ‘Autistic Doctors International’. We also thank Dr David Hillebrandt and Karen Leneh Buckle for their assistance during this project. We are grateful to AsIAm, Ireland’s National Autism Charity and Scally’s SuperValu, Clonakilty, for the funding to enable open access publication.

---

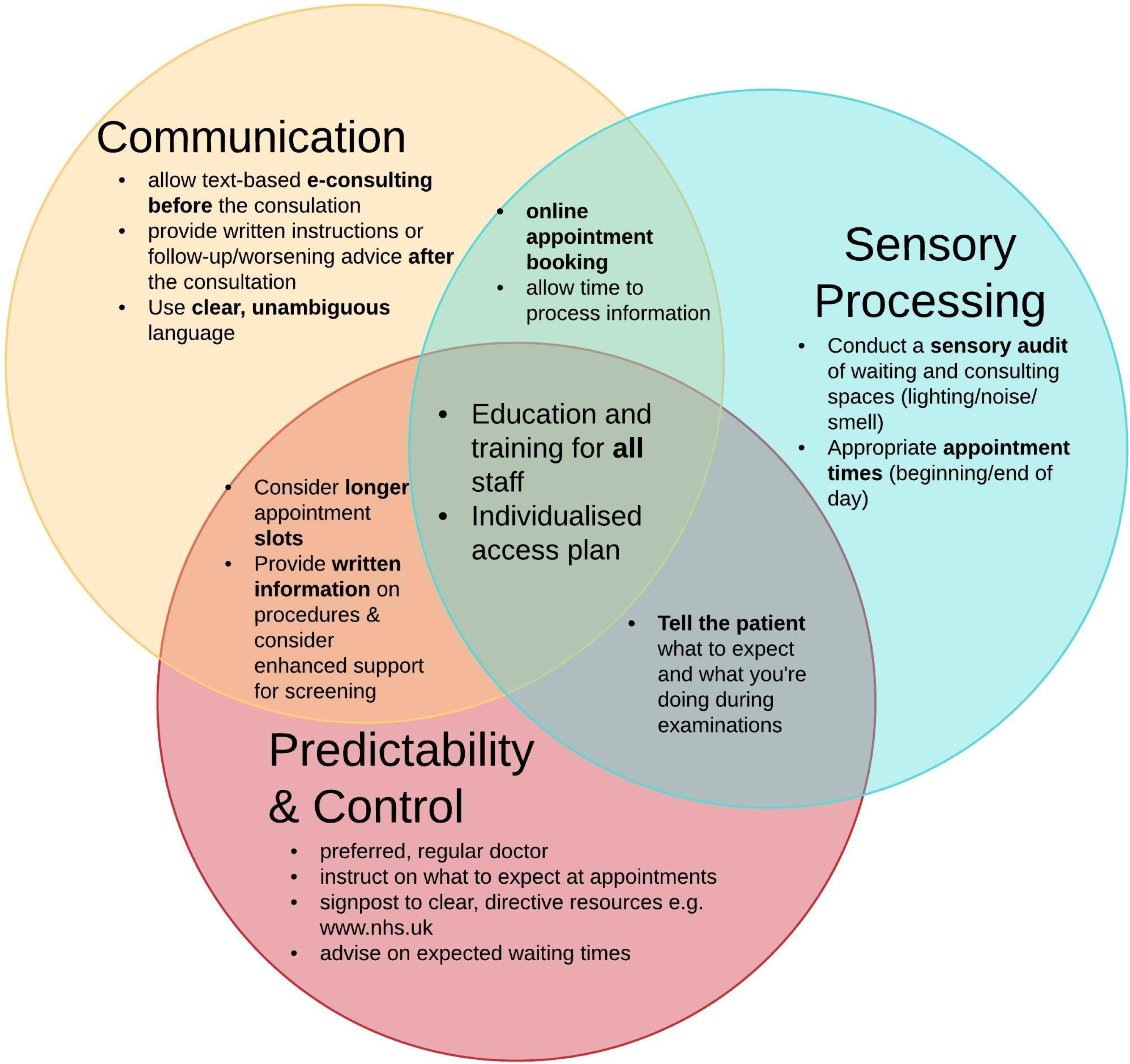

