## Supplemental Tables S1 - S5 for "Barriers to healthcare for autistic adults: Consequences & policy implications. A cross-sectional study": Supplementary Tables.pdf

### Table S1: Predictability & Control

| **Difficulty not knowing:** | **Autistic** | **Parents** | **Control** | **Autistic.Control** | **Autistic.Parent** | **Parent.Control** |
|---|---|---|---|---|---|---|
| Which doctor you will see | 292 (57.6%) | 54 (27.6%) | 38 (24.2%) | p<0.001 ** | p<0.001 ** | p=0.555 ns |
| How long you will wait | 355 (70.0%) | 96 (49.0%) | 47 (29.9%) | p<0.001 ** | p<0.001 ** | p<0.001 ** |
| How long the consultation will last | 201 (39.6%) | 27 (13.8%) | 12 (7.6%) | p<0.001 ** | p<0.001 ** | p=0.098 ns |
| What will happen during the consultation | 320 (63.1%) | 33 (16.8%) | 25 (15.9%) | p<0.001 ** | p<0.001 ** | p=0.932 ns |
| None of the above | 36 (7.1%) | 64 (32.7%) | 84 (53.5%) | p<0.001 ** | p<0.001 ** | p<0.001 ** |

### Table S2: Judgements & Attitudes

| **Going to the doctor, I am anxious :** | **Autistic** | **Parents** | **Control** | **Autistic.Control** | **Autistic.Parent** | **Parent.Control** |
|---|---|---|---|---|---|---|
| I won't be taken seriously when I describe my symptoms | 341 (67.3%) | 51 (26.0%) | 54 (34.4%) | p<0.001 ** | p<0.001 ** | p=0.111 ns |
| I might be wasting the doctor's time | 333 (65.7%) | 71 (36.2%) | 53 (33.8%) | p<0.001 ** | p<0.001 ** | p=0.711 ns |
| I might be considered a hypochondriac | 330 (65.1%) | 43 (21.9%) | 42 (26.8%) | p<0.001 ** | p<0.001 ** | p=0.355 ns |
| about asking for help | 318 (62.7%) | 41 (20.9%) | 28 (17.8%) | p<0.001 ** | p<0.001 ** | p=0.554 ns |
| about discussing mental health | 301 (59.4%) | 57 (29.1%) | 37 (23.6%) | p<0.001 ** | p<0.001 ** | p=0.297 ns |
| that there might be something wrong | 217 (42.8%) | 41 (20.9%) | 55 (35.0%) | p=0.102 ns | p<0.001 ** | p=0.004 ** |
| I don't feel anxious going to the doctor | 17 (3.4%) | 56 (28.6%) | 52 (33.1%) | p<0.001 ** | p<0.001 ** | p=0.420 ns |
| **Is stimming a problem for you at the doctors' office?** | **Autistic** | **Parents** | **Control** | **Autistic.Control** | **Autistic.Parent** | **Parent.Control** |
| Unusual behaviours or stimming elicit negative reactions from other patients | 74 (14.6%) | 69 (35.2%) | 4 (2.5%) | p<0.001 ** | p<0.001 ** | p<0.001 ** |
| Unusual behaviours or stimming elicit negative reactions from other reception staff | 44 (8.7%) | 26 (13.3%) | 1 (0.6%) | p=0.001 ** | p=0.093 ns | p<0.001 ** |
| Unusual behaviours or stimming elicit negative reactions from other medical staff | 37 (7.3%) | 18 (9.2%) | 2 (1.3%) | p=0.009 ** | p=0.498 ns | p=0.003 ** |
| I feel comfortable with stimming at the doctors' office | 101 (19.9%) | 69 (35.2%) | 74 (47.1%) | p<0.001 ** | p<0.001 ** | p=0.031 * |
| I do not feel comfortable with stimming at the doctors' office | 4 (0.8%) | 38 (19.4%) | 5 (3.2%) | p=0.061 ns | p<0.001 ** | p<0.001 ** |
| No need to stim at the doctors' office | 133 (26.2%) | 0 (0.0%) | 0 (0.0%) | p<0.001 ** | p<0.001 ** | |
| I don't understand the term "stimming" | 29 (5.7%) | 11 (5.6%) | 62 (39.5%) | p<0.001 ** | p=1.000 ns | p<0.001 ** |

### Table S3: Planning & Organising

| **Planning & organising** | **Autistic** | **Parents** | **Control** | **Autistic.Control** | **Autistic.Parent** | **Parent.Control** |
|---|---|---|---|---|---|---|
| I find it difficult to prioritise when describing my medical problems | 333 (65.7%) | 38 (19.4%) | 34 (21.7%) | p<0.001 ** | p<0.001 ** | p=0.695 ns |
| I need to give the whole story and not leave anything out | 332 (65.5%) | 30 (15.3%) | 18 (11.5%) | p<0.001 ** | p<0.001 ** | p=0.373 ns |
| I find it difficult to make appointments in advance | 300 (59.2%) | 71 (36.2%) | 45 (28.7%) | p<0.001 ** | p<0.001 ** | p=0.165 ns |
| Making changes to my lifestyle or habits is difficult for me | 282 (55.6%) | 50 (25.5%) | 36 (22.9%) | p<0.001 ** | p<0.001 ** | p=0.663 ns |
| I have forgotten to attend a medical appointment | 230 (45.4%) | 54 (27.6%) | 34 (21.7%) | p<0.001 ** | p<0.001 ** | p=0.251 ns |
| I need to write things down | 227 (44.8%) | 19 (9.7%) | 17 (10.8%) | p<0.001 ** | p<0.001 ** | p=0.863 ns |
| I find waiting difficult | 221 (43.6%) | 26 (13.3%) | 15 (9.6%) | p<0.001 ** | p<0.001 ** | p=0.361 ns |
| I have difficulty making decisions about my health | 222 (43.8%) | 18 (9.2%) | 20 (12.7%) | p<0.001 ** | p<0.001 ** | p=0.369 ns |
| I have turned up for a medical appointment on the wrong day | 151 (29.8%) | 27 (13.8%) | 16 (10.2%) | p<0.001 ** | p<0.001 ** | p=0.390 ns |
| It is difficult to arrange someone to come with me | 103 (20.3%) | 6 (3.1%) | 2 (1.3%) | p<0.001 ** | p<0.001 ** | p=0.446 ns |
| I have forgotten why I made the appointment | 54 (10.7%) | 4 (2.0%) | 1 (0.6%) | p<0.001 ** | p<0.001 ** | p=0.512 ns |
| None of the above | 13 (2.6%) | 53 (27.0%) | 64 (40.8%) | p<0.001 ** | p<0.001 ** | p=0.009 ** |

### Table S4: Support needs

| **Do you visit your doctor:** | **Autistic** | **Parents** | **Control** | **Autistic.Control** | **Autistic.Parent** | **Parent.Control** |
|---|---|---|---|---|---|---|
| Alone, by choice | 306 (60.4%) | 105 (53.6%) | 134 (85.4%) | p<0.001 ** | p=0.121 ns | p<0.001 ** |
| Alone, but would prefer to have a support person | 165 (32.5%) | 3 (1.5%) | 10 (6.4%) | p<0.001 ** | p<0.001 ** | p=0.034 * |
| With a parent, partner or support person | 137 (27.0%) | 15 (7.7%) | 9 (5.7%) | p<0.001 ** | p<0.001 ** | p=0.617 ns |
| With a parent, partner or support person but I would prefer to go alone | 12 (2.4%) | 0 (0.0%) | 0 (0.0%) | p=0.109 ns | p=0.065 ns | |
| With a support animal | 5 (1.0%) | 1 (0.5%) | 0 (0.0%) | p=0.471 ns | p=0.874 ns | p=1.000 ns |
| To support an autistic adult | 26 (5.1%) | 29 (14.8%) | 0 (0.0%) | p=0.008 ** | p<0.001 ** | p<0.001 ** |
| As a parent to access healthcare for my child | 78 (15.4%) | 115 (58.7%) | 30 (19.1%) | p=0.327 ns | p<0.001 ** | p<0.001 ** |
| With my child, but I would prefer to go alone | 6 (1.2%) | 61 (31.1%) | 12 (7.6%) | p<0.001 ** | p<0.001 ** | p<0.001 ** |

**If you were suddenly admitted to hospital, who would be able to bring your personal belongings to you?**

| | Autistic | Parents | Control | Autistic.Control | Autistic.Parent | Parent.Control |
|---|---|---|---|---|---|---|
| Spouse or partner | 234 (46.2%) | 137 (69.9%) | 106 (67.5%) | p<0.001 ** | p<0.001 ** | p=0.715 ns |
| Parent | 168 (33.1%) | 76 (38.8%) | 70 (44.6%) | p=0.012 * | p=0.187 ns | p=0.321 ns |
| Other family member | 96 (18.9%) | 57 (29.1%) | 72 (45.9%) | p<0.001 ** | p=0.005 ** | p=0.002 ** |
| Friend | 118 (23.3%) | 41 (20.9%) | 57 (36.3%) | p=0.002 ** | p=0.569 ns | p=0.002 ** |
| Neighbour | 17 (3.4%) | 6 (3.1%) | 14 (8.9%) | p=0.008 ** | p=1.000 ns | p=0.033 * |
| Paid support person or carer | 15 (3.0%) | 7 (3.6%) | 1 (0.6%) | p=0.174 ns | p=0.860 ns | p=0.139 ns |
| Volunteer support person or carer | 6 (1.2%) | 0 (0.0%) | 0 (0.0%) | p=0.375 ns | p=0.284 ns | |
| Nobody available | 88 (17.4%) | 13 (6.6%) | 5 (3.2%) | p<0.001 ** | p<0.001 ** | p=0.222 ns |

**If you were admitted to hospital for a day case surgical procedure, who would be available to collect you afterwards?**

| | Autistic | Parents | Control | Autistic.Control | Autistic.Parent | Parent.Control |
|---|---|---|---|---|---|---|
| Spouse or partner | 208 (41.0%) | 135 (68.9%) | 100 (63.7%) | p<0.001 ** | p<0.001 ** | p=0.362 ns |
| Parent | 176 (34.7%) | 78 (39.8%) | 73 (46.5%) | p=0.010 * | p=0.242 ns | p=0.248 ns |
| Other family member | 96 (18.9%) | 60 (30.6%) | 75 (47.8%) | p<0.001 ** | p=0.001 ** | p=0.001 ** |
| Friend | 116 (22.9%) | 46 (23.5%) | 67 (42.7%) | p<0.001 ** | p=0.947 ns | p<0.001 ** |
| Neighbour | 17 (3.4%) | 4 (2.0%) | 8 (5.1%) | p=0.446 ns | p=0.503 ns | p=0.201 ns |
| Paid support person or carer | 17 (3.4%) | 9 (4.6%) | 0 (0.0%) | p=0.042 * | p=0.577 ns | p=0.017 * |
| Volunteer support person or carer | 3 (0.6%) | 0 (0.0%) | 0 (0.0%) | p=0.776 ns | p=0.664 ns | |
| Nobody available | 99 (19.5%) | 14 (7.1%) | 3 (1.9%) | p<0.001 ** | p<0.001 ** | p=0.042 * |

**If you needed assistance at home after an operation, who would be available to provide that care?**

| | Autistic | Parents | Control | Autistic.Control | Autistic.Parent | Parent.Control |
|---|---|---|---|---|---|---|
| Spouse or partner | 219 (43.2%) | 123 (62.8%) | 101 (64.3%) | p<0.001 ** | p<0.001 ** | p=0.846 ns |
| Parent | 152 (30.0%) | 71 (36.2%) | 74 (47.1%) | p<0.001 ** | p=0.132 ns | p=0.050 * |
| Other family member | 83 (16.4%) | 50 (25.5%) | 61 (38.9%) | p<0.001 ** | p=0.008 ** | p=0.010 * |
| Friend | 74 (14.6%) | 31 (15.8%) | 43 (27.4%) | p<0.001 ** | p=0.772 ns | (p=0.012 * |

| | | | | | | |
|---|---|---|---|---|---|---|
| Neighbour | 14 (2.8%) | 7 (3.6%) | 8 (5.1%) | p=0.241 ns | p=0.750 ns | p=0.660 ns |
| Paid support person or carer | 30 (5.9%) | 11 (5.6%) | 3 (1.9%) | p=0.071 ns | p=1.000 ns | p=0.135 ns |
| Volunteer support person or carer | 4 (0.8%) | 0 (0.0%) | 0 (0.0%) | p=0.599 ns | p=0.491 ns | |
| Nobody available | 131 (25.8%) | 34 (17.3%) | 13 (8.3%) | p<0.001 ** | p=0.022 * | p=0.020 * |

**If you are a parent and you were unable to care for your child due to illness, who would be available to provide that care to your child?**

| | Autistic | Parents | Control | Autistic.Control | Autistic.Parent | Parent.Control |
|---|---|---|---|---|---|---|
| Spouse or partner | 110 (21.7%) | 135 (68.9%) | 64 (40.8%) | p<0.001 ** | p<0.001 ** | p<0.001 ** |
| Parent | 37 (7.3%) | 63 (32.1%) | 40 (25.5%) | p<0.001 ** | p<0.001 ** | p=0.211 ns |
| Other family member | 40 (7.9%) | 52 (26.5%) | 40 (25.5%) | p<0.001 ** | p<0.001 ** | p=0.919 ns |
| Friend | 21 (4.1%) | 24 (12.2%) | 27 (17.2%) | p<0.001 ** | p<0.001 ** | p=0.245 ns |
| Neighbour | 4 (0.8%) | 8 (4.1%) | 8 (5.1%) | p<0.001 ** | p=0.007 ** | p=0.843 ns |
| Paid support person or carer | 5 (1.0%) | 20 (10.2%) | 9 (5.7%) | p<0.001 ** | p<0.001 ** | p=0.185 ns |
| Volunteer support person or carer | 1 (0.2%) | 1 (0.5%) | 0 (0.0%) | p=1.000 ns | p=1.000 ns | p=1.000 ns |
| Nobody available | 34 (6.7%) | 27 (13.8%) | 6 (3.8%) | p=0.256 ns | p=0.005 ** | p=0.003 ** |
| I don't have a child requiring care | 274 (54.0%) | 1 (0.5%) | 78 (49.7%) | p=0.387 ns | p<0.001 ** | p<0.001 ** |

### Table S5: Facilitators

| **Visits to my doctor would be easier if:** | **Autistic** | **Parents** | **Control** | **Autistic.Control** | **Autistic.Parent** | **Parent.Control** |
|---|---|---|---|---|---|---|
| I could book an appointment online | 339 (66.9%) | 113 (57.7%) | 102 (65.0%) | p=0.732 ns | p=0.028 * | p=0.197 ns |
| I could email my doctor in advance with a description of the issue I need to discuss | 316 (62.3%) | 51 (26.0%) | 35 (22.3%) | p<0.001 ** | p<0.001 ** | p=0.493 ns |
| I could wait in a quiet place or outside until it was my turn | 284 (56.0%) | 45 (23.0%) | 13 (8.3%) | p<0.001 ** | p<0.001 ** | p<0.001 ** |
| I could book the first or last appointment of the day | 210 (41.4%) | 79 (40.3%) | 38 (24.2%) | p<0.001 ** | p=0.854 ns | p=0.002 ** |
| I could book an appointment by text | 209 (41.2%) | 54 (27.6%) | 44 (28.0%) | p=0.004 ** | p=0.001 ** | p=1.000 ns |
| There was a sensory box available in the waiting room | 80 (15.8%) | 28 (14.3%) | 4 (2.5%) | p<0.001 ** | p=0.707 ns | p<0.001 ** |
| None of the above | 16 (3.2%) | 32 (16.3%) | 34 (21.7%) | p<0.001 ** | p<0.001 ** | p=0.255 ns |
